# Health Disparities and Reporting Gaps in Artificial Intelligence (AI) Enabled Medical Devices: A Scoping Review of 692 U.S. Food and Drug Administration (FDA) 510k Approvals

**DOI:** 10.1101/2024.05.20.24307582

**Authors:** Vijaytha Muralidharan, Boluwatife Adeleye Adewale, Caroline J Huang, Mfon Thelma Nta, Peter Oluwaduyilemi Ademiju, Pirunthan Pathmarajah, Man Kien Hang, Oluwafolajimi Adesanya, Ridwanullah Olamide Abdullateef, Abdulhammed Opeyemi Babatunde, Abdulquddus Ajibade, Sonia Onyeka, Zhou Ran Cai, Roxana Daneshjou, Tobi Olatunji

## Abstract

Machine learning and artificial intelligence (AI/ML) models in healthcare may exacerbate health biases. Regulatory oversight is critical in evaluating the safety and effectiveness of AI/ML devices in clinical settings. We conducted a scoping review on the 692 FDA 510k-approved AI/ML-enabled medical devices to examine transparency, safety reporting, and sociodemographic representation. Only 3.6% of approvals reported race/ethnicity, 99.1% provided no socioeconomic data. 81.6% did not report the age of study subjects. Only 46.1% provided comprehensive detailed results of performance studies; only 1.9% included a link to a scientific publication with safety and efficacy data. Only 9.0% contained a prospective study for post-market surveillance. Despite the growing number of market-approved medical devices, our data shows that FDA reporting data remains inconsistent. Demographic and socioeconomic characteristics are underreported, exacerbating the risk of algorithmic bias and health disparity.

## Introduction

To date, the FDA has approved 692 medical devices driven by artificial intelligence and machine learning (AI/ML) for potential use in clinical settings. (1) Most recently, the FDA has launched the Medical Device Development Tools (MDDT) program, which aims to “facilitate device development, timely evaluation of medical devices and promote innovation”, with the Apple Watch being the first approved device for this regulatory process (2,3). As AI/ML studies begin to translate to clinical environments, it is crucial that end users can evaluate the applicability of devices to their unique clinical settings and assess sources of bias and risk.

One definition of algorithmic bias in the context of AI/ML health systems is instances when an algorithm amplifies inequities and results in poor healthcare outcomes. (4) Some defined sub-categories of algorithmic bias are listed below. (5)

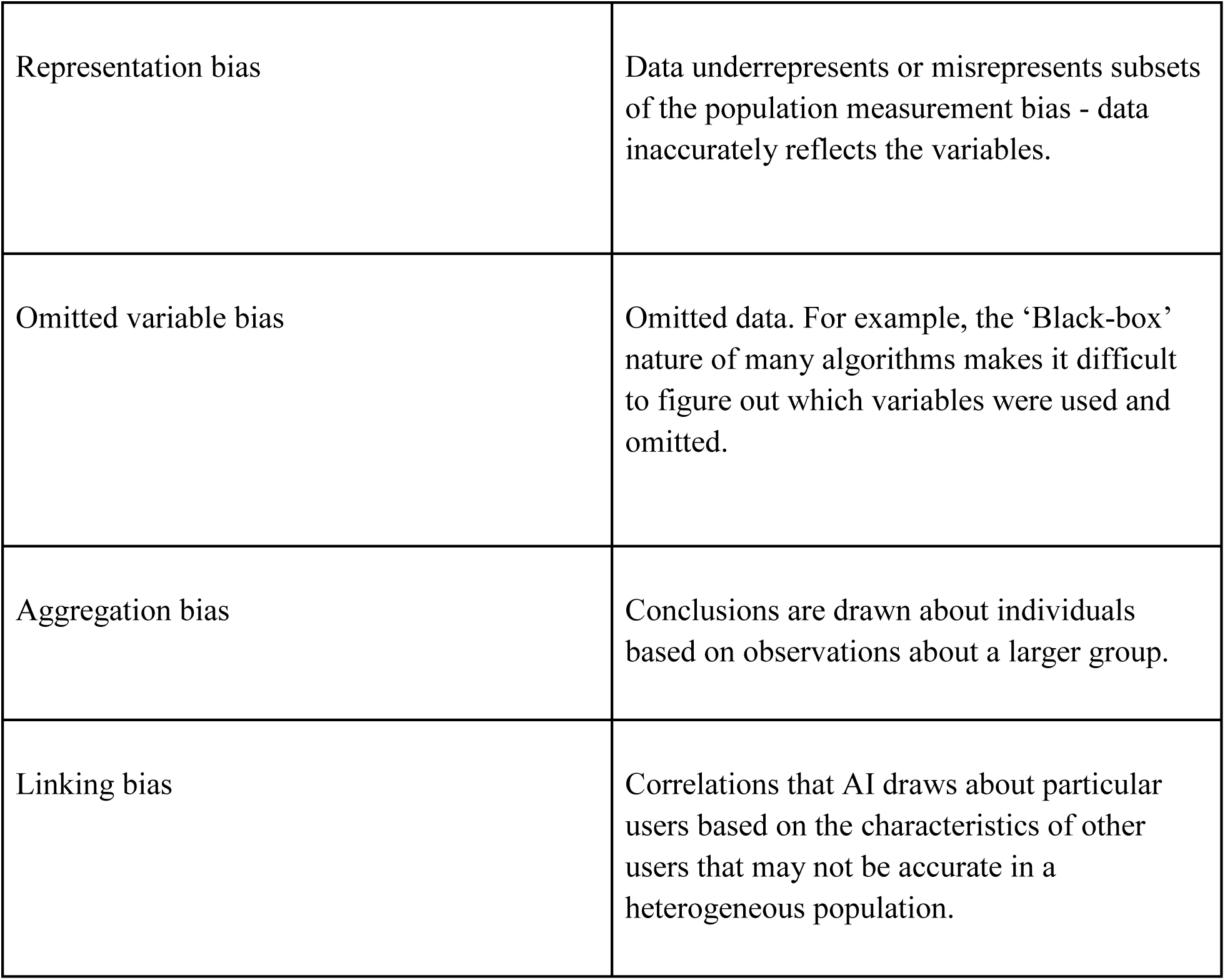

|  |  |
| --- | --- |
| Representation bias | Data underrepresents or misrepresents subsets of the population measurement bias - data inaccurately reflects the variables. |
| Omitted variable bias | Omitted data. For example, the ‘Black-box’ nature of many algorithms makes it difficult to figure out which variables were used and omitted. |
| Aggregation bias | Conclusions are drawn about individuals based on observations about a larger group. |
| Linking bias | Correlations that AI draws about particular users based on the characteristics of other users that may not be accurate in a heterogeneous population. |

Despite the rise in awareness of algorithmic bias and its potential implications on the generalizability of AI/ML models (6), there is a paucity of standardized data reporting by regulatory bodies including the FDA that provide reliable and consistent information on the development, testing, and training of algorithms for clinical use. This limits accurate analysis and evaluation of algorithmic performance, particularly in the context of under-represented research groups such as ethnic minorities, children, maternal health patients, patients with rare diseases, and those from lower socioeconomic strata. Deploying devices that cannot be transparently evaluated by end users may increase health disparity and is particularly relevant in the context of emerging clinical trials and real-world deployment. (7)

Here, we investigate AI-as-medical-device Food and Drug Administration (FDA) approvals to examine the contents, consistency, and transparency in FDA reporting of market-approved devices with a focus on bias.

## Methods

We conducted a scoping review of AI-as-a-medical-device approved by the FDA between 1995 and 2023, using FDA Summary of Safety and Effectiveness Data (SSED). (8) SSED is a public document made available following approval of a device by the FDA. FDA approval is conditioned upon the determination that probable benefits outweigh probable risks as well as reasonable evidence of safety and effectiveness. (9)

All SSEDs of FDA-approved AI/ML-enabled medical devices between 1995 and 2023 made available at the <u>FDA Artificial Intelligence and Machine Learning (AI/ML)-Enabled Medical</u> <u>Devices page</u> were included. (1) Each SSED was reviewed by an expert in computer science, medicine, or academic clinical research who identified, extracted, and entered relevant variables of interest (Supplementary Table 1). Data was then computed into a Microsoft Excel spreadsheet (Supplementary Table 2).

Variables of interest were determined per the Consolidated Standards of Reporting Trials - Artificial Intelligence (CONSORT-AI) extension checklist which is a guideline developed by international stakeholders to promote transparency and completeness in reporting AI clinical trials. (10) Equivocal or unclear information identified in each SSED was then evaluated in consensus.

Primary outcome measures included frequency of race/ethnicity reporting, age reporting, and availability of sociodemographic data of the algorithmic testing population provided in each approval document. Secondary outcomes evaluated the representation of various medical specialties, organ systems, and specific patient populations such as pediatric and geriatric in approved devices.

## Results

### Distribution of device approval across clinical specialties

692 SSEDs of FDA-approved AI-enabled medical devices/software were analyzed. There was a steady increase in annual FDA approvals for AI-enabled medical devices with a mean of 7 between 1995 and 2015, increasing to 139 approvals in 2022 (Figure 1). The regulatory class of each device included in the study was determined and categorized according to the United States Food and Drug Administration (FDA) classification system. Only 2 (0.3%) of the devices belonged to the regulatory Class III, while the vast majority (99.7%) of the devices belonged to Class II.

**Figure 1:**
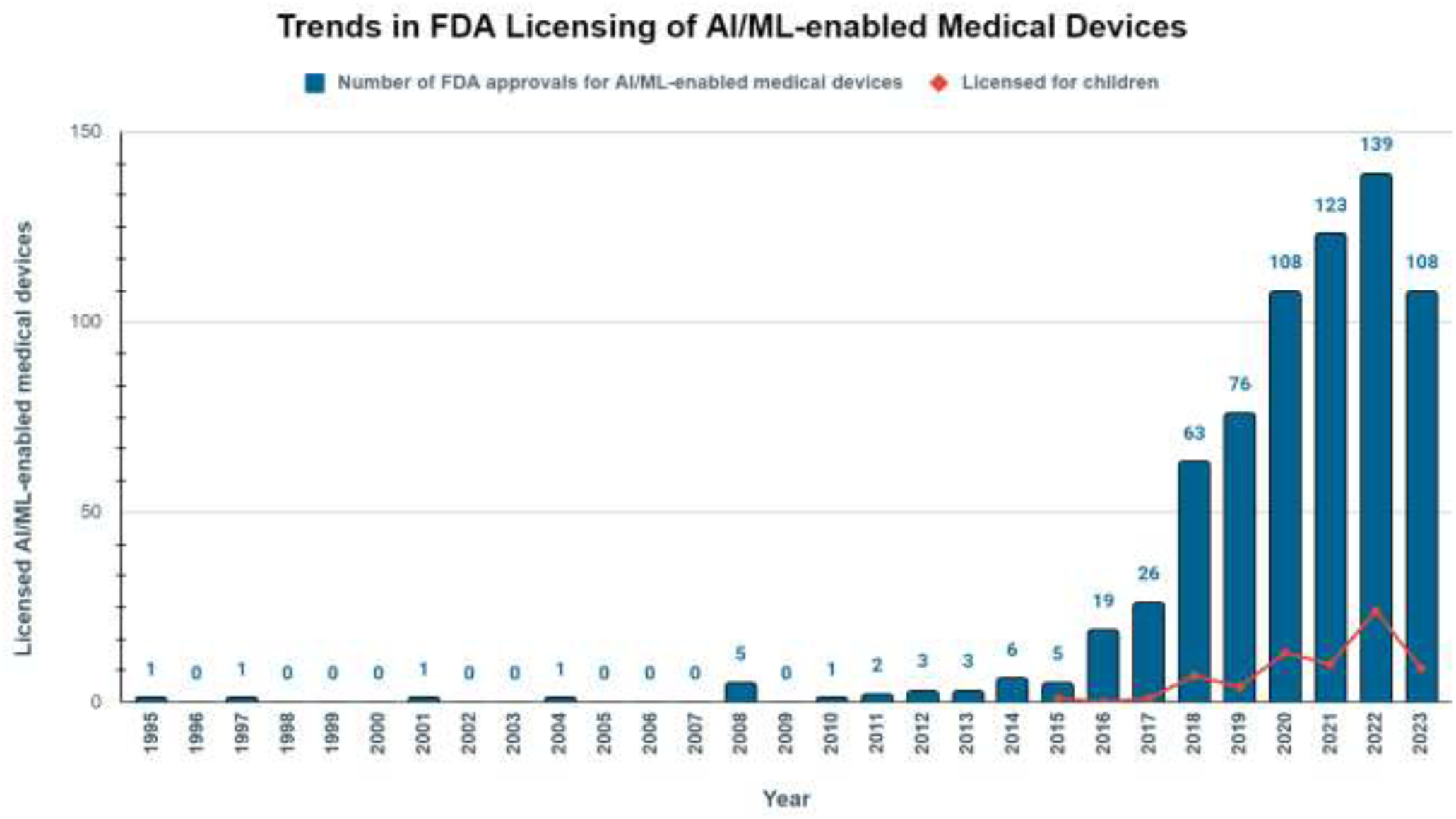
Trends in FDA Licensing of AI/ML-enabled Medical Devices.

Table 1 shows the distribution of 408 approved devices across organ systems. The top three organ systems represented amongst approved medical devices are the circulatory (20.8%), nervous (13.6%), and reproductive (7.2%). The least represented are the urinary (1.2%) and endocrine (0.7%) systems (Table 1). Each device in the FDA database is classified under a particular medical specialty (Figure 2). The FDA classification shows that the most represented medical specialty is Radiology (532 approvals; 76.9%) with the fewest approvals in Immunology, Orthopedics, Dental Health, Obstetrics, and Gynecology (Figure 2, Table 2). A total of 284 (40.1%) approved devices could not be categorized to an organ system either because (I) the clinical indication was not specific to one system or because (II) the function of the device cuts across multiple organ systems (e.g. whole-body imaging system/software). As such, there are some differences between the categories of organ system and medical specialty. For instance, 70 (10.1%) of the devices are classified by the FDA under the cardiovascular field despite 144 (20.8%) approvals specific to the circulatory system (Table 1, Table 2).

**Figure 2:**
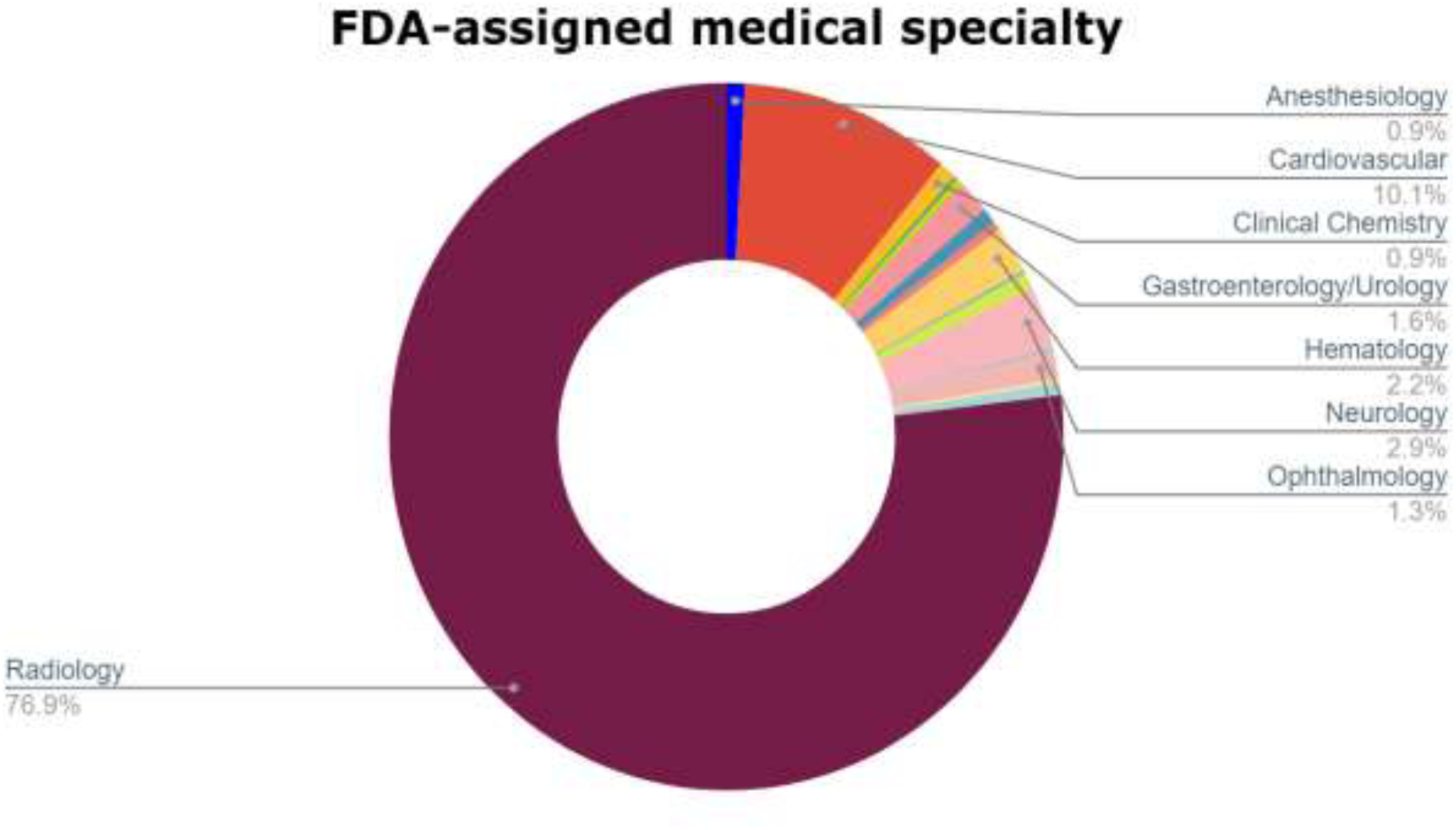
FDA-assigned medical specialty.

**Table 1:** Distribution of FDA-approved AI-enabled medical devices by organ system.

| System | Number of approvals (%) |
| --- | --- |
| Circulatory | 144 (20.8) |
| Nervous | 94 (13.6) |
| Respiratory | 48 (6.9) |
| Reproductive | 47 (6.8) |
| Musculoskeletal | 33 (4.8) |
| Gastrointestinal | 24 (3.5) |
| Urinary | 8 (1.2) |
| Hematologic | 5 (0.7) |
| Endocrine | 5 (0.7) |

**Table 2:** Primary medical specialty associated with FDA approval.

| Medical Specialty | Number of Approval (%) |
| --- | --- |
| Radiology | 532 (76.9) |
| Cardiovascular | 70 (10.1) |
| Neurology | 20 (2.9) |
| Hematology | 15 (2.2) |
| Gastroenterology/Urology | 11 (1.6) |
| Ophthalmology | 9 (1.3) |
| Anesthesiology | 6 (0.9) |
| Clinical Chemistry | 6 (0.9) |
| General And Plastic Surgery | 5 (0.7) |
| Microbiology | 5 (0.7) |
| General Hospital | 3 (0.4) |
| Ear Nose & Throat | 2 (0.3) |
| Pathology | 4 (0.6) |
| Dental | 1 (0.1) |
| Immunology | 1 (0.1) |
| Obstetrics And Gynecology | 1 (0.1) |
| Orthopedic | 1 (0.1) |

### Reporting data on statistical parameters and post-market surveillance

Indication for use of the device was reported in most (678; 98.0%) SSEDs (Figure 3a) and 487 (70.4%) SSEDs contained a pre-approval performance study. However, 435 (62.8%) provided no data on the sample size of the subjects. Although 319 (46.1%) provided comprehensive detailed results of performance studies including statistical analysis, only 13 (1.9%) of them included a link to a scientific publication with further information on the safety and efficacy of the device (Figure 4). Only 219 (31.6%) SSEDs provided data on the underlying machine-learning technique. Only 62 device documents (9.0%) contained a prospective study for post-market surveillance. 14 (2.0%) SSEDs addressed reporting of potential adverse effects of medical devices on users.

**Figure 3:**
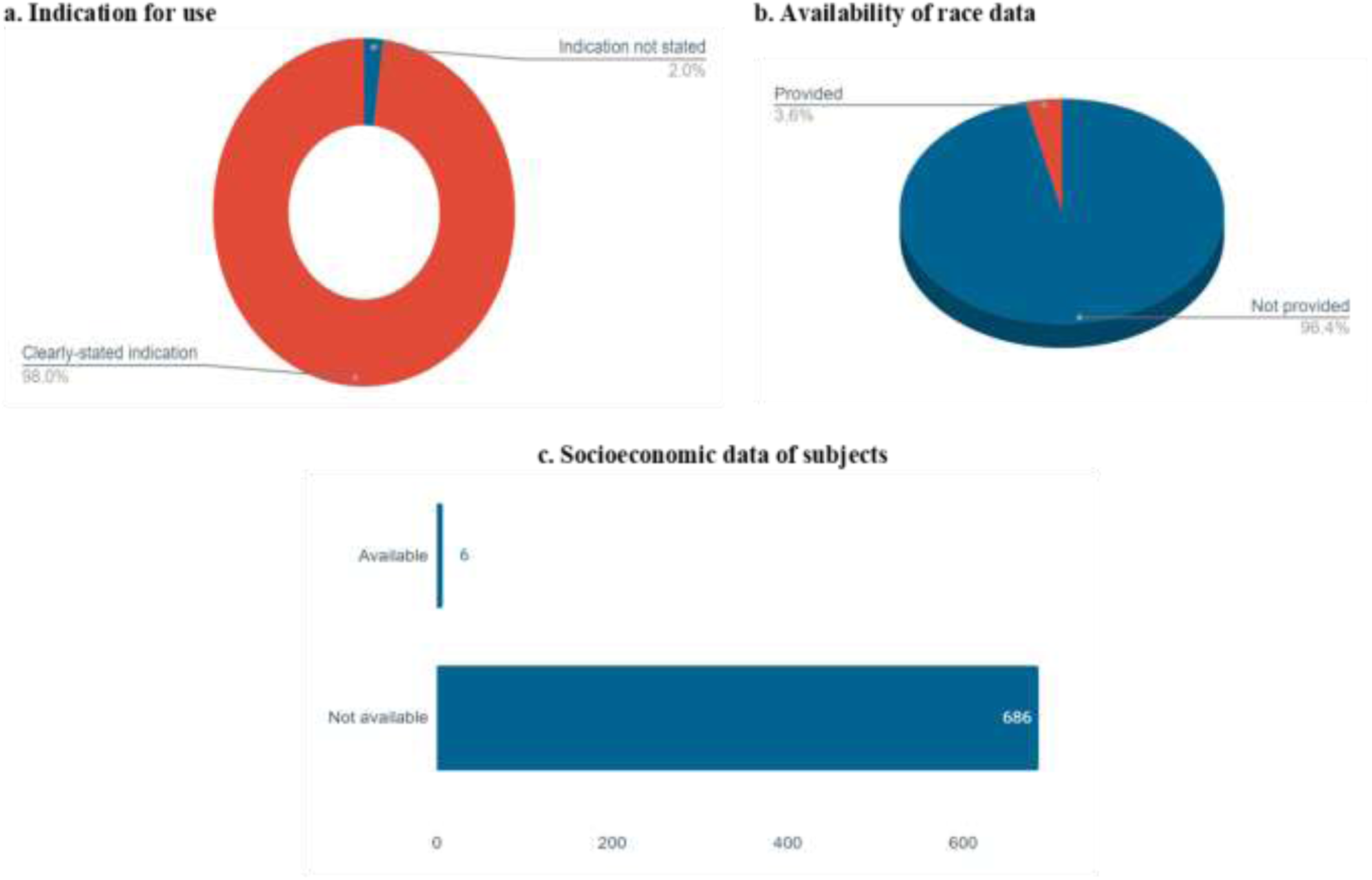
Reported Information on Intended Subjects.

**Figure 4:**
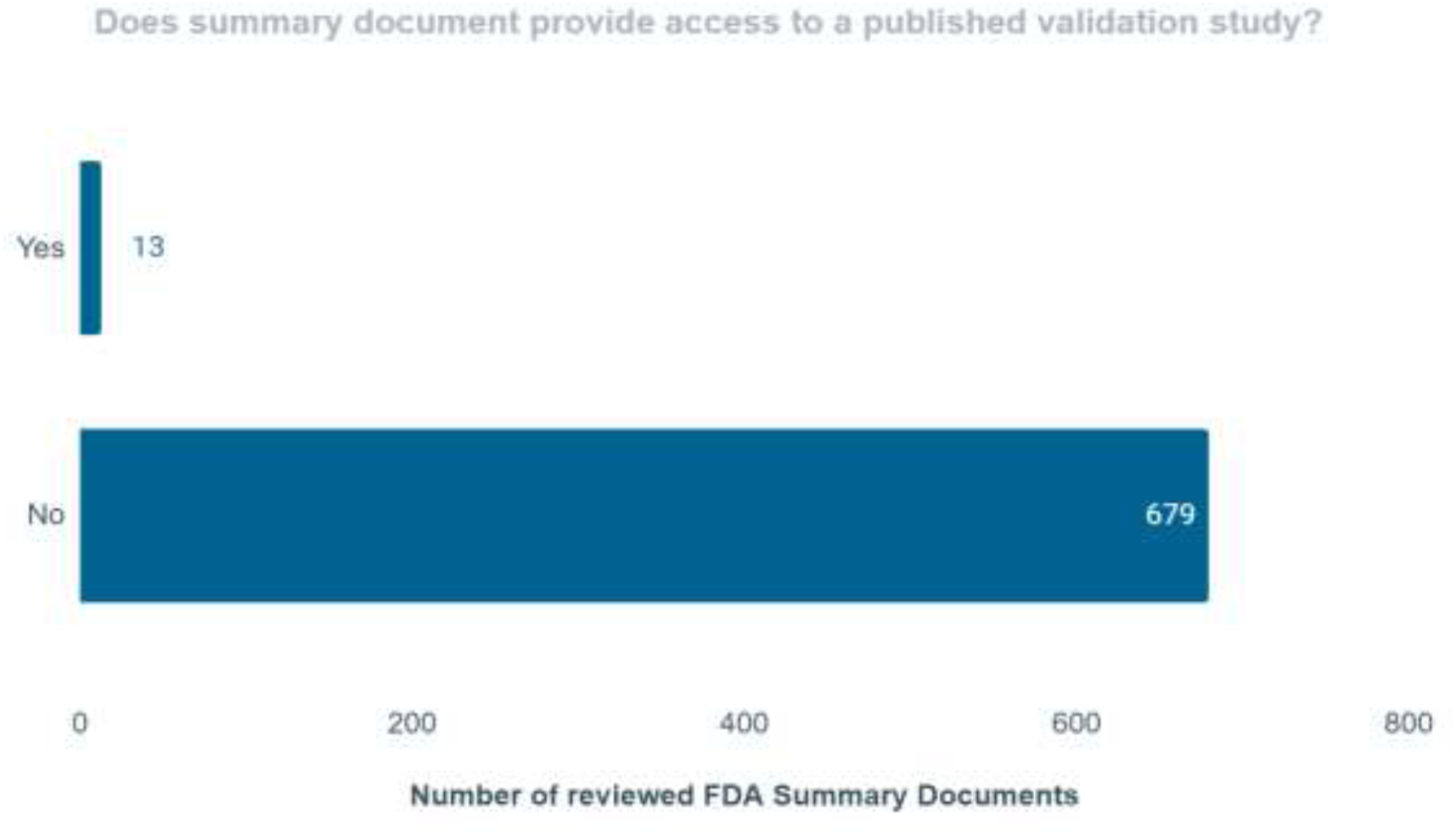
Accessibility to publications supporting safety, efficacy, and transparency.

### Race, ethnicity, and socioeconomic diversity

Patient demographics in algorithmic testing data were only specified in 153 (22.1%) SSEDs, with 539 (77.9%) not providing any demographic data (Figure 3b). Only 25 (3.6%) provided information on the race and/or ethnicity of tested or intended users. Socioeconomic data on tested or intended users were provided for only 6 (0.9%) of devices (Figure 3c).

### Age Diversity

There were 134 (19.4%) SEEDs with available information on the age of the intended subjects. Upon examining age diversity in approved devices, the first FDA approval for a device licensed for children was in 2015. Between 2015 and 2022, the annual FDA approvals for the pediatric age group steadily increased from 1 to 24 in total. Despite this rise, the proportion of pediatric-specific approvals relative to the total approvals (for adults and pediatrics combined) has remained low, fluctuating between 0.0% and 20.0% (Figure 1, Table 3). Although 4 (0.6%) devices were exclusively developed for children, we found sixty-five more devices that have been approved for use in both adult and pediatric populations, thus bringing the total number of approvals for the pediatric population to 69 (10.0%). Testing and validation of devices in children and adults was reported in only 134 (19.4%) SSEDs (Figures 5a, 5b). The distribution of devices for children (n = 69) across medical specialties falls under just 5 categories, following a similar pattern as earlier observed for the entire population with lead representation in the fields of Radiology (72.5%; n = 69), Cardiovascular health (14.4%; n = 69) and Neurology (10.1%; n = 69) (Figure 5c). There were only three (0.4%) approved devices that focused exclusively on geriatric health.

**Figure 5:**
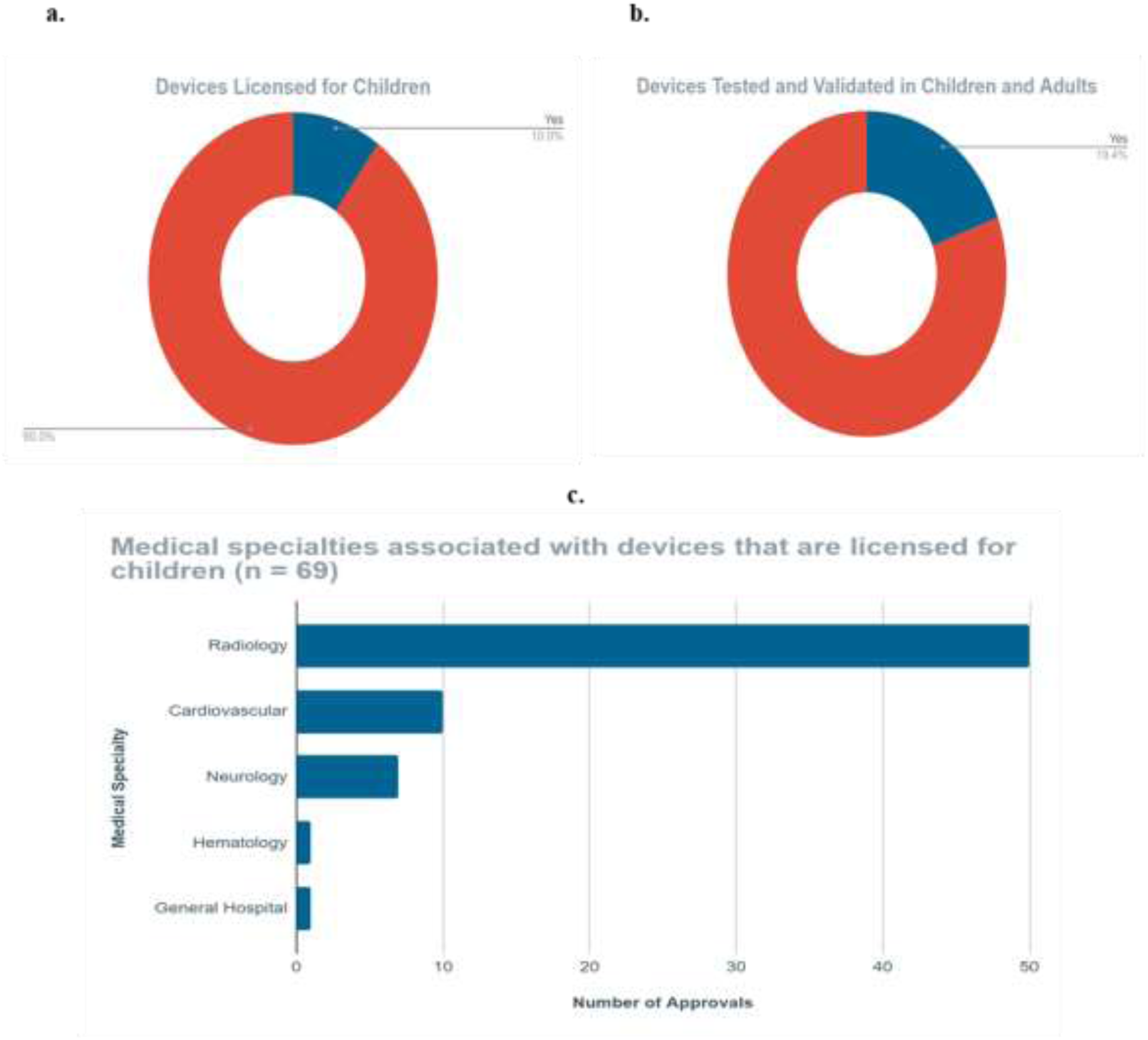
FDA Approvals for Pediatric-Licensed Devices.

**Table 3:** Annual trends in FDA approvals for AI-enabled medical devices and software for children. (**2015–2023**)

| **Year** | **Number of FDA approvals for AI/ML-enabled medical devices** | **Licensed for children (percentage relative to total FDA approvals %)** |
| --- | --- | --- |
| 2015 | 5 | 1 (20.0) |
| 2016 | 19 | 0 (0.0) |
| 2017 | 26 | 1 (3.8) |
| 2018 | 63 | 7 (11.1) |
| 2019 | 76 | 4 (5.3) |
| 2020 | 108 | 13 (12.0) |
| 2021 | 123 | 10 (8.1) |
| 2022 | 139 | 24 (17.3) |
| 2023 | 108 | 9 (8.3) |

### Gender Diversity

When examining gender reporting transparency, there were a total of 39 (5.6%) approvals exclusively for women’s health, 36 of them focusing on the detection of breast pathology. The remaining three were designed to aid cervical cytology; determine the number and sizes of ovarian follicles; and perform fetal/obstetrics ultrasound. Of the 10 (1.5%) devices that were exclusively for men, eight of them were indicated in diagnostic and/or therapeutic procedures involving the prostate, while the remaining two were for seminal fluid analysis.

## Discussion

Our study highlights a lack of consistency and data transparency in published FDA AI/ML approval documents which may exacerbate health disparities. In a similar study examining 130 FDA-approved AI medical devices between January 2015 and December 2020, 97% reported only retrospective evaluations; prospective studies did not evaluate high-risk devices; 72% did not publicly report whether the algorithm was tested on more than one site; and 45% did not report basic descriptive data such as sample size. (11,12) A lack of consistent reporting prevents objective analysis of the fairness, validity, generalizability, and applicability of devices for end users. As our results describe, only 37% of device approval documents contained information on sample size. As the clinical utility of algorithmic data is limited by data quantity and quality (13), a lack of transparency in sample size reporting significantly limits the accurate assessment of the validity of performance studies, and device effectiveness. (14)

Only 14.5% of devices provided race or ethnicity data. Recent literature strongly emphasizes the risks of increasing racial health disparity through the propagation of algorithmic bias. (15–17) A lack of racial and ethnic profiling in publicly available regulatory documents risks further exacerbating this important health issue. (18,19) The FDA has recognized the potential for bias in AI/ML-based medical devices and has initiated action plans (“Artificial Intelligence/Machine Learning (AI/ML)-Based Software as a Medical Device (SaMD) Action Plan”) in January 2021 (20,21) to address these concerns. However, despite these efforts, our study highlights reporting inconsistencies that may continue to propagate racial health disparities. (22) In light of these results, there is a pressing need for transparent and standardized regulatory frameworks that explicitly consider racial diversity in the evaluation and reporting of AI/ML medical devices. (23) Other strategies to mitigate racial bias may include adopting adversarial training frameworks and implementing post-authorization monitoring to ensure AI/ML devices perform equitably across all patient demographics. (24,25)

While AI/ML presents potential opportunities to reduce socioeconomic disparity in health, a lack of representation of target users across varied economic strata risks the propagation of health disparity in higher and lower-income groups. (26) As with other clinical research domains, a lack of representation of lower socioeconomic groups including those in remote and rural areas, risks neglect of those most likely to benefit from improved access to healthcare. (27,28) Our data shows that only 0.6% of approved devices contained specific data detailing the socioeconomic striate of users in testing and/or algorithmic training datasets. This data renders it difficult to predict the potential clinical and financial impacts of approved medical devices on economic population subsets. Furthermore, a lack of socioeconomic data prevents accurate and robust cost-effectiveness analyses that may significantly impact the availability and impact of medical algorithms or devices. (29,30). Studies have underscored disparities rooted in socioeconomic factors, impacting the performance of AI/ML technologies. (4,31,32) Initiatives promoting diversity in data collection and consideration of model performance across socioeconomic groups are paramount and must be incorporated in the assessment of market approval for emerging technologies. (33)

With only 19.4% of devices providing information on the age of intended device users, our study suggests that the evaluation and approval process of medical AI devices by the FDA lacks comprehensive data on age diversity. Recent literature across specialties demonstrates differential performances in algorithms trained on adult or pediatric data. (34,35) As an example, a study exploring echocardiogram image analysis suggested that adult images could not be appropriately generalized to pediatric patients and vice versa. (36) A lack of transparent age reporting, therefore, risks propagating age-related algorithmic bias, with potential clinical, ethical, and societal implications on the target population. (34,37) Mitigating age bias requires a concerted effort to ensure that training and testing datasets appropriately match intended users.

Further, with only 0.6% of devices approved specifically for the pediatric age group, our findings identify equity gaps in the representation of children in AI/ML market-approved devices. (38,39)

With our findings showing that only 0.4% of approved devices cater specifically to geriatric health needs, specific considerations should be considered for the older adult population. Despite having the highest proportion of healthcare utilization, geriatric patients are traditionally underrepresented in clinical research. (34,40,41) A recent WHO ethical guidance document outlines the potential societal, clinical, and ethical implications of ageism in medical research, and describes the lack of geriatric representation as a health hazard in light of aging populations. (42,43) Initiatives such as the NIH’s Inclusion Across the Lifespan policy aim to promote the participation of older adults in research studies, which may help equitize the potential impacts of algorithmic development for this population, considering unique ethical and clinical considerations. (44,45) Similar to considerations for children, we propose that regulatory bodies encourage market approval documents to make clear intentions to test and train on a geriatric population and ensure that appropriate validation methods are in place to ensure the appropriate generalization of model outputs to specific geriatric health needs. (46,47)

Our study also examined variations in the representation of different medical specialties among approved medical devices. Specialties most commonly represented include Radiology, Cardiology, and Neurology. (1) Promoting clinical equity requires a more balanced representation of specialties and disease systems in digital innovation. Whilst we appreciate that AI/ML research is limited by data availability and data quality, industry, academia, and clinicians must advocate for equality of innovation amongst specialties, to include a broad range of conditions and patient populations in medical device development and testing that may potentially benefit. (48) As the FDA is a US-based regulator, our review does not examine the representation of specialties or conditions outside the US, and in particular in Low- and Middle-Income Countries (LMICs) which contain over 80% of the global burden of disease. (49–51)

Many countries do not have the regulatory capacity to release approval documentation, and thus future studies must incorporate international data availability, collaboration, and cohesion. (52) Regulatory bodies both within and outside the USA must attempt to align technological development with key priorities in national and global disease burden to promote global equity. (53)

Our results showed that transparency in study enrollment, study design methodology, statistical data, and model performance data were significantly inconsistent amongst approved devices. While 70.4% of studies provided some detail on performance studies before market approval, only 46.1% provided detailed results of the performance studies. In 62.9% of devices, there was no information provided on sample size. Transparency is crucial in addressing the challenges of interpretability and explainability in AI/ML systems, and our current findings suggest that evaluation cannot be comprehensively conducted across approved FDA devices. (54) Models that are transparent in their decision-making process (interpretability) or those that can be elucidated by secondary models (explainability) are essential for validating the clinical relevance of any outcomes and ensuring that devices that may be incorporated in clinical settings and thus enhanced transparency must be incorporated in future approvals. (23,55) Further ethical considerations encompass a range of issues, including patient privacy, consent, fairness, accountability, and algorithmic transparency. (56) Including ethics methods in both study protocols and future regulatory documents may minimize privacy concerns arising from the potential misuse, and increase end-user confidence. (57,58)

Only 142 (20.5%) of the reviewed devices provide statements on potential risks to end users. Further, only 13 (1.9%) approval documents included a corresponding published scientific validation study, providing evidence of their safety and effectiveness. Underreporting of safety data in approved devices limits the ability of end users to determine generalizability, effectiveness, cost-effectiveness, and medico-legal complexities that may occur from device incorporation. (59) It is therefore paramount that regulatory bodies such as the FDA advocate for a mandatory release of safety data and considerations of potential adverse events. One example of an approved device reporting adverse effects is the Brainomix 360 e-ASPECTS, a computer-aided diagnosis (CADx) software device used to assist the clinician in the characterization of brain tissue abnormalities using CT image data. (60) Its safety report highlights some of the potential risks of incorrect scoring of the algorithm, the potential misuse of the device to analyze images from an unintended patient population, and device failure.

Here, we detail some recommendations that may be adopted by the FDA and similar regulatory bodies internationally to reduce the risk of bias and health disparity in AI/ML.

Recommendations for regulatory reporting of AI/ML devices
|  |  |
| --- | --- |
| Explicit Representation Standards | The FDA should explicitly mandate that AI/ML medical device submissions provide comprehensive demographic information, including race, ethnicity, age, gender, and socioeconomic status of the studied populations. This ensures that the algorithms are developed and validated on diverse datasets, minimizing the risk of biased outcomes and ensuring equitable representation. |
| Inclusive Validation Criteria | Establish clear guidelines for validation studies to include diverse and representative populations. Manufacturers should be required to demonstrate the performance of their AI/ML algorithms across various demographic groups, ensuring that the technology is effective for minorities, marginalized, and underrepresented communities. |
| Bias Mitigation Protocols | Enforce the inclusion of explicit methodologies for identifying and mitigating biases in AI/ML algorithms. Manufacturers should be required to document their efforts to address algorithmic biases throughout the development process, ensuring that the final product minimizes disparities in health outcomes. |
| Transparency Requirements | Implement stringent transparency standards, mandating clear |
|  | documentation of the machine learning techniques used, the sources of training data, and the rationale behind algorithmic decisions. Transparency ensures that end-users, healthcare providers, and regulatory authorities have insight into the algorithm's decision-making processes. |
| Post-Market Surveillance for Equity | Establish a robust post-market surveillance framework specifically focused on monitoring the performance of AI/ML devices in real-world settings. This ongoing evaluation should include an analysis of outcomes across diverse patient populations, with a particular emphasis on detecting and addressing any emerging biases affecting minorities and under-represented groups. |
| Interdisciplinary Review Panels | Form interdisciplinary review panels within the FDA consisting of experts in data science, healthcare disparities, and ethics. This ensures a holistic evaluation of AI/ML submissions, incorporating perspectives that can identify and address potential biases and disparities in health outcomes. |
| Incentives for Diversity and Equity | Establish incentives, such as expedited review processes or regulatory benefits, for manufacturers who proactively address issues of representation, validation rigor, and bias mitigation in their AI/ML device submissions. This encourages |
|  | industry-wide commitment to fairness and equity. |

While the FDA’s Artificial Intelligence/Machine Learning (AI/ML) Action Plan outlines steps to advance the development and oversight of AI/ML-based medical devices (21), including initiatives to improve transparency, post-market surveillance, and real-world performance monitoring, (61) our study highlights that there remain several clinically relevant inconsistencies in market approval data that may exacerbate algorithmic biases and health disparity.

The ramifications of inadequate demographic, socioeconomic, and statistical information in the majority of 501(k) submissions to the FDA for AI/ML medical devices approved for clinical use are multifaceted and extend across societal, health, legal, and ethical dimensions (10,34,62).

Addressing these informational gaps is imperative to ensure the responsible and equitable integration of AI/ML technologies into clinical settings and the appropriate evaluation of demographic metrics in clinical trials. Additional focus must be given to under-represented groups who are most vulnerable to health disparities as a consequence of algorithmic bias. (34,63)

## Data Availability

All data produced in the present work are contained in the manuscript

## Author Contributions

VM, BAA, RD and TO were involved in conceptualization, study design and methodology. VM, BAA, CJH, MN, PA, PP, MKH, OA, ROA, AOB, AA, SO and ZRC were responsible for screening data sources, data extraction and entry. BAA and TO provided formal data analysis.

VM, BAA, CJH, RD and TO ensured data accuracy. VM, BAA, CJH, MN, PA and PP, MKH and AOB wrote the original draft of the manuscript. VM, BAA, CJH, MN, PA and PP revised and wrote the final draft. RD and TO revised the final draft and provided supervision for the project.

## Author Contributions

The authors declare no competing interests.

